# Cortical thickness and cognitive decline in Parkinson patients with deep brain stimulation in the MARK-PD study

**DOI:** 10.1101/2023.05.16.23290035

**Authors:** Robert Schulz, Merve Uyar, José A. Graterol Perez, Monika Pötter-Nerger, Carsten Buhmann, Christian Gerloff, Chi-un Choe

## Abstract

Advanced Parkinson’s disease (PD) and deep brain stimulation (DBS) are associated with cognitive impairment. We aimed to evaluate whether regional cortical thickness is associated with cognitive decline in patients with advanced PD treated with DBS. From the MARK-PD study, 32 patients with DBS implantation were included. Cortical thickness of 148 brain areas and cognitive function were assessed with MRI data using the FreeSurfer pipeline and Montreal Cognitive Assessment at baseline and follow-up (median 17 months), respectively. Adjusted linear and Cox regression models were calculated. Thinner orbitofrontal, cingulate, and occipito-temporal cortices as well as cuneus is related to significant cognitive decline. Moreover, lower thickness of these cortices is also associated with faster cognitive decline. Our study suggests that PD patients treated with DBS and lower cortical thickness in multiple brain regions develop at stronger and faster cognitive decline during the later course of the disease.

## Introduction

Structural brain imaging in Parkinson’s disease (PD) has contributed to our understanding of how cortical brain regions undergo structural alterations and to what extent these changes are associated with cognitive decline, a common and disabling feature of advanced PD ^1,2^. Deep brain stimulation (DBS) frequently utilized in more impaired patients carries an additional risk of treatment-induced cognitive decline ^3^. For PD in general, evidence from cross-sectional ^4–8^ and longitudinal studies ^9–15^ underlines that reduced cortical thickness (CT) or grey matter volume occur in orbitofrontal, occipito-temporal, parietal and cingulate cortices, which are associated with cognitive decline. For patients treated with DBS, studies have related electrode positions to odds of DBS induced cognitive decline. For instance, electrode trajectories intersecting with caudate nuclei increase the risk of a decline after surgery in global cognition and working memory performance ^16^. More recent network analyses showed that stimulation sites affecting structural brain networks involving the caudate nuclei, orbitofrontal, cingulate, parietal cortices, the hippocampus, and cognitive regions of the cerebellum are at particular risk for cognitive decline ^17^. However, it remains unclear if measures of cortical anatomy of such brain regions operationalized by CT might relate to cognitive decline in PD patients treated with DBS. Therefore, we have longitudinally evaluated cognitive assessments of PD patients who were already treated with DBS from the observational MARK-PD cohort study.^15^ Structural MRI data collected before DBS and study inclusion were integrated into regression models. We hypothesized that lower CT values of orbitofrontal, cingulate, and occipito-temporal cortices relate to future cognitive decline in PD patients treated with DBS.

## Participants and methods

### Participants and clinical assessment

The Biomarkers in Parkinson’s disease (MARK-PD) study is a prospective observational single-centre study with biobanking at the University Medical Centre Hamburg-Eppendorf ^18^. For the current study, we have included 32 PD patients who had undergone DBS treatment in the past. Cognitive assessment was performed at *baseline* (timepoint T_1_) and *follow-up* (timepoint T_2_). The study protocol was approved by the local ethics committee (PV5298). The investigation was conducted in accordance with the Declaration of Helsinki. Written informed consent was obtained from all participants. Clinical assessment was performed as previously described ^18^. Cognitive assessment (Montreal Cognitive Assessment, MoCA) was performed in the medication and stimulation ON state. Past medical history and medication were documented from medical records.

### Imaging acquisition & cortical thickness analysis

Clinical data were integrated with retrospective available structural MRI data collected for routine clinical purposes, before DBS, in the past (median 399, range 959 – 14 days) before T_1_. The datasets were obtained from two different scanners: 31 patients were scanned with a 3T Skyra scanner (Siemens, Erlangen, Germany) applying a 3-dimensional magnetization-prepared rapid gradient echo (3D-MPRAGE) sequence (repetition time (TR)=1900 ms, echo time (TE)=2.46 ms, inversion time (TI)=900 ms, flip angle 9°, field of view (FOV)=240 mm, voxel dimension of 0.94 mm^3^). One patient was scanned using a 3T Ingenia scanner (Philips, Amsterdam, Netherlands) using a standard 3D T1-weighted protocol with a TR of 8.04 – 8.10 ms, TE of 3.69 – 3.7 ms, flip angle 8°, FOV 238 - 248 mm and a voxel dimension of 0.94×0.94×1 mm. Datasets were processed with FreeSurfer version 7.1.0 (http://surfer.nmr.mgh.harvard.edu/) using the default options to measure CT. The reconstructions were visually inspected and, if required, manually corrected following established recommendations from the FreeSurfer’s documentation. Surface data was smoothed with a full-width-half-maximum Gaussian kernel of 10 mm. Mean CT values were finally calculated over 148 anatomical labels including gyri and sulci using the Destrieux cortical parcellation ^20^.

### Statistical analysis

Statistical analyses were carried out in R version 4.0.3 (R Core Team, 2020; r-project.org) or with IBM SPSS Statistics (Version 27, IBM Corp., Armonk, NY). Linear regression analyses were used to correlate regional CT values with change in MoCA between T_1_ and T_2_. As previously shown ^19^ we have splitted the patient group by the CT median in groups with thinner and thicker CT than the median. Change of MoCA between T_1_ and T_2_ was treated as the dependent variable, dichotomized binary CT was treated as the independent variable of interest. The target effect CT was corrected for age, disease duration, time interval between MRI scan and T_1_ (MRI-T_1_), time interval T_1_-T_2_ and MoCA at T_1_. Results are given as mean differences between groups (with 95% confidence intervals, CI) with values > 0 indicating increased decline in MoCA in patients exhibiting lower CT values compared to patients with higher values. The independent association between CT and time-to-cognitive decline, defined as a decrease of more than 2 points in the MoCA score during the observational period ^18^, was determined by multivariable Cox regression analyses. Results are given as hazard ratios with 95% confidence intervals (CI), regressions were adjusted for age, disease duration, MRI-T_1_ and MoCA at T_1_. Kaplan-Meier curves were used for illustration. Statistical significance was assumed at *P* value < 0.05. Fsbrain package in R was used for illustrations.

## Results

### Baseline characteristics and cognitive decline

For this study, 32 PD patients with DBS before study inclusion were analysed. These patients were on average 63.5 ± 9.1 years old, 20 were male, MDS-UPDRS III scores were (median [interquartile range]) 20 [15, 26], Hoehn&Yahr stages were 2 [2, 3] and disease duration was 13 [9, 15] years (Table 1). Follow-up time was 529 [341, 728] days. The MoCA score at T_1_ was 27 [25, 29] and at T_2_ 25.5 [22, 28].

**Table 1.** Demographic and clinical characteristics. Data are mean (SD), n (%) or median [IQR], as appropriate. Categorical variables are given as numbers (percentages) of participants. Abbreviations LED, L-dopa equivalence dose; MoCA, Montreal cognitive assessment; MDS-UPDRS, Movement Disorders Society-Unified Parkinson’s disease Ranking Scale.

| Characteristics | T<sub>1</sub> |  |
| --- | --- | --- |
| Age, years | 63.5 ± 9.1 |  |
| Male | 20 |  |
| Disease duration, years | 13 [9, 15] |  |
| Education, days | 13.5 [12.25, 14.5] |  |
| Hypertension, % | 12 |  |
| Diabetes, % | 1 |  |
| Prior stroke, % | 1 |  |
| Time T<sub>1</sub> to MRI, days | -399 [-273, -716] |  |
| LED, mg/d | 801 [499, 1000] |  |
| Follow-up, days | 529 [341, 728] |  |
|  | T<sub>1</sub> | T<sub>2</sub> |
| UPDRS III, points | 20 [15, 26] | 27 [17, 30] |
| Hoehn&Yahr, stage | 2 [2, 3] | 2.5 [2, 3] |
| MoCA, points | 27 [25, 29] | 25.5 [22, 28] |

### Cortical thickness and cognitive decline

Lower CT values in selected orbitofrontal, cingulate, and occipito-temporal cortices were significantly related to larger cognitive decline of MoCA scores between T_1_ and T_2_ (Figure 1). Specifically, patients with thinner cortices in regions summarized by Table 2 exhibited an absolute decrease in MoCA score from T_1_ to T_2_ of up to 3.9 points. Conversely, patients with higher CT values did not exhibit any significant cognitive decline during the observational period. Absolute decrease in MoCA scores ranged between 3.9 and 3.4 (*P*<0.05 for all 9 regions) in patients with thinner cortices. For patients with higher CT, values ranged between 0.7 and 0.2 points (not significant). Importantly, patients allocated to groups according to CT values below and above the median did not differ in MoCA at T_1_ scoring at the beginning of the observational period. Moreover, none of the covariates evaluated reached statistical significance in the models.

**Figure 1.**
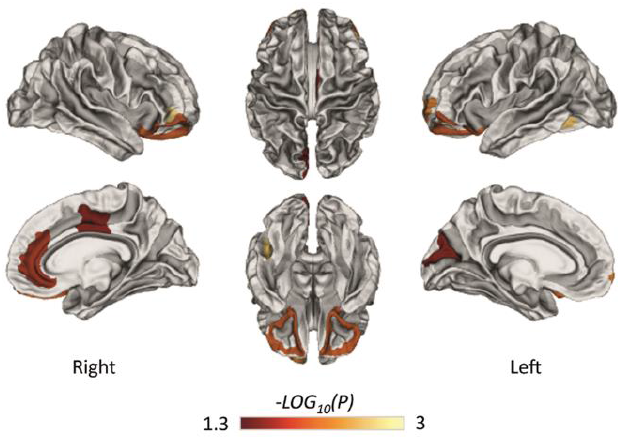
Regional CT and extend of cognitive decline. Brain regions exhibiting significant associations between CT and change in MoCA during the observational period are illustrated with -LOG_10_(*P)* values color-coded. Right and left hemispheres are given. Upper row shows lateral and top views, lower row shows views from the mid sagittal plane and from the bottom.

**Table 2.**
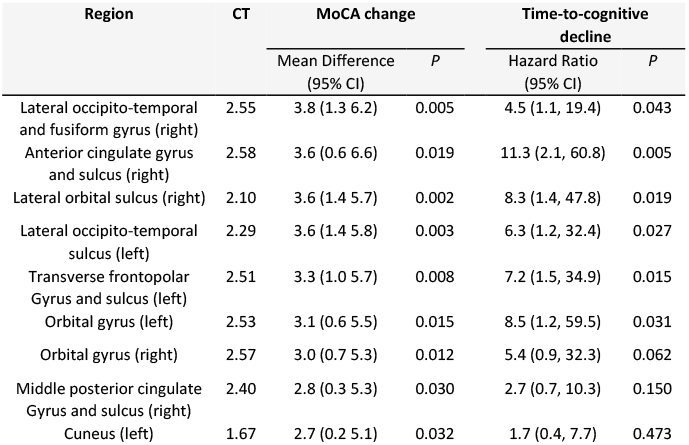
Regional cortical thickness associates with cognitive decline. Linear regression analysis of MoCA change (mean difference with 95% confidence intervals, CI) and Cox regression analysis for time-to-cognitive decline (hazard ratio (HR) with 95% CI) are given for patients with lower CT values compared to patients with higher values (reference). *P* values are uncorrected. CT indicates CT median values (in mm) used for group allocation.

| Region | CT | MoCA change |  | Time-to-cognitive decline |  |
| --- | --- | --- | --- | --- | --- |
|  |  | Mean Difference (95% CI) | *P* | Hazard Ratio (95% CI) | *P* |
| Lateral occipito-temporal and fusiform gyrus (right) | 2.55 | 3.8 (1.3 6.2) | 0.005 | 4.5 (1.1, 19.4) | 0.043 |
| Anterior cingulate gyrus and sulcus (right) | 2.58 | 3.6 (0.6 6.6) | 0.019 | 11.3 (2.1, 60.8) | 0.005 |
| Lateral orbital sulcus (right) | 2.10 | 3.6 (1.4 5.7) | 0.002 | 8.3 (1.4, 47.8) | 0.019 |
| Lateral occipito-temporal sulcus (left) | 2.29 | 3.6 (1.4 5.8) | 0.003 | 6.3 (1.2, 32.4) | 0.027 |
| Transverse frontopolar Gyrus and sulcus (left) | 2.51 | 3.3 (1.0 5.7) | 0.008 | 7.2 (1.5, 34.9) | 0.015 |
| Orbital gyrus (left) | 2.53 | 3.1 (0.6 5.5) | 0.015 | 8.5 (1.2, 59.5) | 0.031 |
| Orbital gyrus (right) | 2.57 | 3.0 (0.7 5.3) | 0.012 | 5.4 (0.9, 32.3) | 0.062 |
| Middle posterior cingulate Gyrus and sulcus (right) | 2.40 | 2.8 (0.3 5.3) | 0.030 | 2.7 (0.7, 10.3) | 0.150 |
| Cuneus (left) | 1.67 | 2.7 (0.2 5.1) | 0.032 | 1.7 (0.4, 7.7) | 0.473 |

### Cortical thickness and time to cognitive decline

Finally, time-to-event analyses were carried out to answer if the CT estimates, which are associated with a larger extent of cognitive decline, are also linked with a faster cognitive decline. In adjusted Cox regression analysis, six of these regions showed a significant relationship between lower CT values at baseline and faster cognitive decline until follow-up (Table 2, Figure 2).

**Figure 2.**
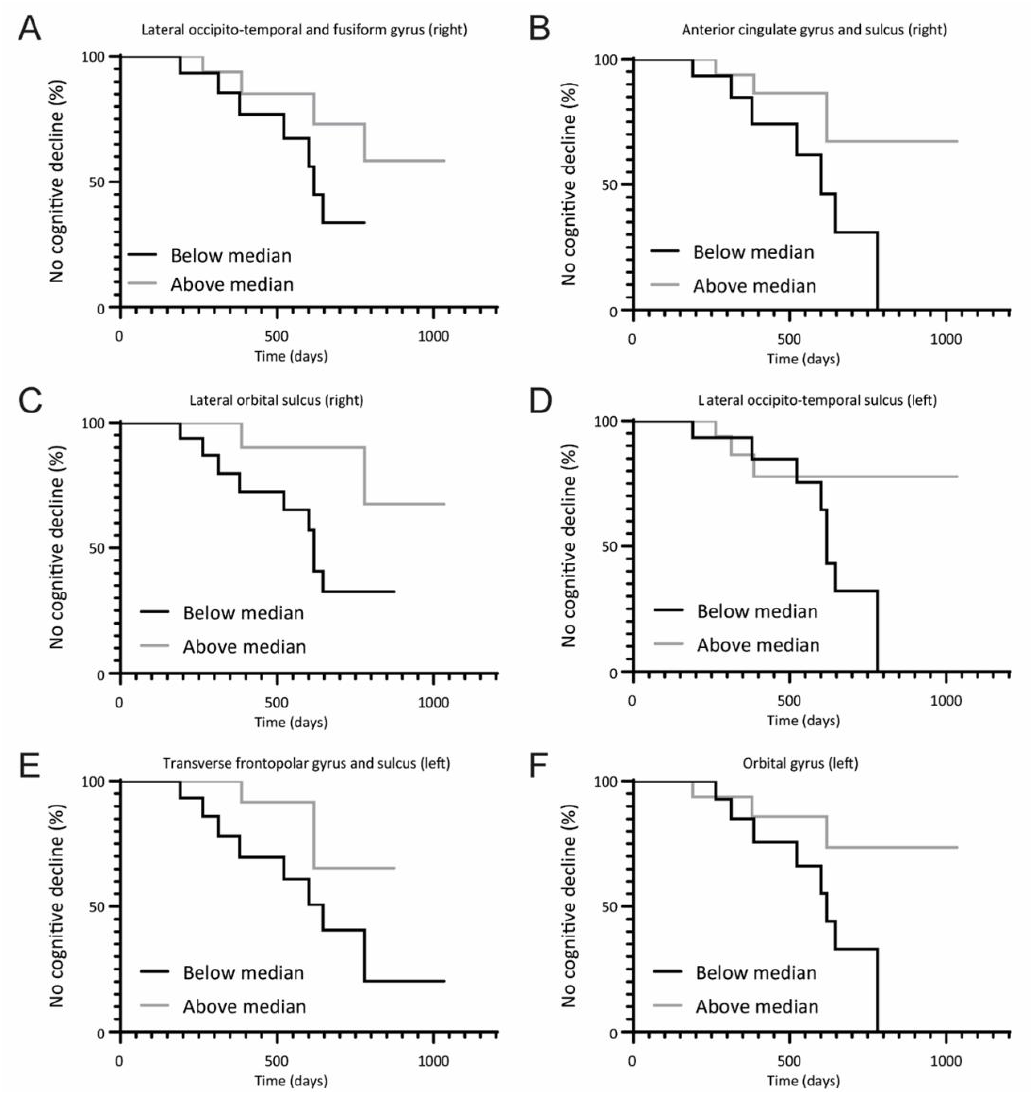
Regional CT and time to cognitive decline. A -F Kaplan-Meier plots illustrating time-to-cognitive decline in PD patients with regional CT below and above the median for six different regions.

## Discussion

The main finding of the present study is that thinner orbitofrontal, cingulate, and occipito-temporal cortices as well as cuneus are related to cognitive decline in PD patients previously implanted with DBS. Interestingly, patients with higher CT values did not exhibit any change in MoCA, whereas patients with lower values showed an absolute decrease of up to 4 points. Moreover, thinner cortices in some of these regions link to faster cognitive decline.

These data complement recent structural network data in PD patients acquired right after DBS of the subthalamic nucleus showing that electrode position and stimulation site critically influence the risk of DBS-induced cognitive decline after surgery. Specifically, stimulation sites connected to the bilateral orbitofrontal cortices and cingulate cortices of the right hemisphere were associated with cognitive decline ^17^. In excellent agreement with previous results, an impaired structural state of orbitofrontal and cingulate cortices was associated with a worse cognitive outcome in DBS treated PD patients. Studies in PD patients without DBS revealed that cortical thinning was observed in regions along the superior frontal gyrus, caudal anterior cingulate cortex, medial occipital and temporal cortices in individuals converting to mild cognitive impairment (MCI) or dementia ^9,10^. Furthermore, PD patients with MCI showed grey matter atrophy in the orbitofrontal cortex, cuneus and fusiform gyrus when compared to patients without MCI_8_. Similarly, PD patients with conversion from MCI to dementia showed CT reductions in orbitofrontal regions. One longitudinal study has directly related the extent of cortical thinning in temporal and medial occipital cortices over time to cognitive decline ^13^. Corroborating these data and accounting particularly for baseline MoCA, age and disease duration, out results argue that preserved structural brain integrity of orbitofrontal, cingulate and occipito-temporal cortices as well as cuneus might attenuate cognitive decline in PD. Our findings are in line with emerging concepts of brain reserve capacity for neurological diseases ^21–23^. We suggest potential cut-off values for CT, which need validation in larger prospective studies. Today, there is still a need to identify patients who are at increased risk of cognitive decline after DBS surgery. In addition to the known clinical parameters such as MoCA scores, CT of orbitofrontal, cingulate, and occipito-temporal cortices as well as cuneus might be a useful complemental predictor in the future to infer the individual patients’
s cognitive risk. For instance, one pre-post DBS analysis in 59 PD patients of similar age, disease duration and Hoehn & Yahr stage showed an average decline in MoCA of only 0.8 within 6 months after DBS ^3^. The present longitudinal analysis of clinical data obtained from advanced PD patients previously implanted with DBS evidenced a decline in MoCA of almost 4 points in patients with lower CT values. This might render CT before DBS particularly helpful in detecting patients at highest risks to develop cognitive decline after DBS.

There are several critical limitations to note. First, MRI data were acquired up to 2.6 years before T_1_. Statistical modelling accounted for this variability in MRI-T_1_. We did not observe any significant influence of this factor for change of MoCA between T_1_ and T_2_. Nevertheless, our results require validation in larger prospective studies comprising clinical and MRI data at one time point. Second, our cohort included patients who had already undergone DBS with variable time intervals between DBS surgery and study inclusion. An appropriate control group of PD patients with similar demographic and disease-specific characteristics is missing. Although the present findings are in very good agreement with previous DBS-specific network data ^17^, we cannot infer any DBS specificity for the present structure-outcome relationships. Third, data of electrode sites, volumes of activated tissue and affected brain networks were not integrated in the present analyses. Such information might be helpful to further investigate whether lower CT in *patients at risk* regarding their network characteristics ^17^ might further aggravate cognitive decline analogous to a *second hit*. Fourth, the observational period showed a large variability (median 17 months, range 6-34 months). However, we adjusted our models for T_1_-T_2,_ which did not influence the results. Fifth, we used CT values as dichotomized binary variables to increase the statistical power and to overcome the limitation of potential outliers and influential points. Group allocation of individual patients based on median split is specific to the present cohort. Thus, the present results might change in external independent datasets. Sixth, *P* values are presented uncorrected for multiple comparisons due to the exploratory design. Finally, the present analyses were focused on CT estimates of cortical anatomy. Information regarding other brain structures such as subcortical nuclei were not considered in this work. A combined modelling of cortical and subcortical brain anatomy would require larger samples to shed novel light on structural determinants of cognitive functioning and decline in PD.

Overall, our study suggests that PD patients previously treated with DBS and thinner orbitofrontal, cingulate and occipito-temporal cortices as well as cuneus develop at stronger and faster cognitive decline during the later course of the disease.

## Data Availability

All data produced in the present study are available upon reasonable request to the authors

## Funding

CUC and RS are supported by an Else Kröner Exzellenzstipendium from the Else Kröner-Fresenius-Stiftung (grant numbers 2018_EKES.04 to CUC, 2020_EKES.16 to RS).

## Conflict of Interest

RS, MU and JGP have nothing to declare. MPN received lecture fees from Abbvie, Abbott and Boston scientific and served as consultant for Medtronic, Boston scientific, Licher, Zambon and Abbvie. CB served on scientific advisory boards for Bial, Desitin, Kyowa Kirin, Merz, STADA Pharm and Zambon and received honoraria for lectures from Abbvie, Bial, Desitin, TAD Pharma, UCB Pharma and Zambon. CG reports personal fees from AMGEN, personal fees from Boehringer Ingelheim, Novartis, Daiichi Sankyo, Abbott, Prediction Biosciences and Bayer. CUC reports personal fees from Pfizer and Zambon.

## Author contributions

RS and CUC designed and organized the project, designed, and performed the statistical analyses and wrote the first draft. MU and JGP executed the project and contributed to the statistical analyses. MPN, CB and CG contributed to the statistical analyses. All authors revised the manuscript for important intellectual content.

## Data availability

The data that support the findings of this study are available on reasonable request from the corresponding author.

## Notes

### Funding Statement

CUC and RS are supported by an Else Kroener Exzellenzstipendium from the Else Kroener-Fresenius-Stiftung (grant numbers 2018_EKES.04 to CUC, 2020_EKES.16 to RS).

### Author Declarations

The study protocol was approved by the local ethics committee (PV5298) of the Chamber of Physicians Hamburg.

